# Progress Towards UNAIDS’s 95-95-95 Targets in Zimbabwe: Sociodemographic Constraints and Geospatial Heterogeneity

**DOI:** 10.1101/2023.07.26.23293207

**Authors:** MD Tuhin Chowdhury, Anna Bershteyn, Masabho Milali, Daniel Citron, Sulani Nyimbili, Godfrey Musuka, Diego F Cuadros

## Abstract

The HIV/AIDS epidemic remains critical in sub-Saharan Africa, with UNAIDS establishing “95-95-95” targets to optimize HIV care. Using the Zimbabwe Population-based HIV Impact Assessment (ZIMPHIA) geospatial data, this study aimed to identify patterns in these targets and determinants impacting the HIV care continuum in underserved Zimbabwean communities. Analysis techniques, including Gaussian kernel interpolation, optimized hotspot, and multivariate geospatial k-means clustering, were utilized to establish spatial patterns and cluster regional HIV care continuum needs. Further, we investigated healthcare availability, access, and social determinants and scrutinized the association between socio-demographic and behavioral covariates with HIV care outcomes. Disparities in progress toward the “95-95-95” targets were noted across different regions, with each target demonstrating unique geographic patterns, resulting in four distinct clusters with specific HIV care needs. Key factors associated with gaps in achieving targets included younger age, male sex, employment, and minority or no religious affiliation. Our study uncovers significant spatial heterogeneity in the HIV care continuum in Zimbabwe, with unique regional patterns in “95-95-95” targets. The spatial analysis of the UNAIDS targets presented here could prove instrumental in designing effective control strategies by identifying vulnerable communities that are falling short of these targets and require intensified efforts. Our result provides insights for designing region-specific interventions and enhancing community-level factors, emphasizing the need to address regional gaps and improve HIV care outcomes in vulnerable communities lagging behind.

## 1 Introduction

HIV is a leading global health priority. In 2021, an estimated 38 million people were living with HIV (PLHIV), 1.5 million new HIV infections, and 650,000 AIDS-related deaths globally [1]. To address this high burden, the Joint United Nations Programme on HIV/AIDS (UNAIDS) has established the 95-95-95 targets, whereby 95% of PLHIV should be diagnosed, 95% of those diagnosed with HIV should be receiving antiretroviral therapy (ART), and 95% of all those receiving ART should achieve viral suppression (VLS). This would result in 85% of all PLHIV with viral load suppression, helping to reduce HIV transmission and mortality [2].

Progress toward the 95-95-95 targets is particularly vital in sub-Saharan Africa – home to two-thirds of PLHIV – because HIV has spread more broadly than in other global populations. In Zimbabwe, HIV is widespread in every geographic area, with 12.9% of all adults infected, ranging from 9.6% of adults in the province of Manicaland to 15.4% of adults in Matabeleland South. To assess progress toward 95-95-95 targets, a population-based HIV impact assessment (PHIA) was conducted in 2016, and again in 2020 [3, 4]. The proportion of PLHIV aware of their HIV status was 76.8% in 2016 and 86.8% in 2020. Of those aware of their status, the proportion on ART was 88.4% in 2016 and 97.0% in 2020. Of those on ART, the proportion with suppressed viral load was 85.3% in 2016 and 90.3% in 2020 [5]. Although these indicators show strong progress on average, it is not clear whether progress has been uniform across Zimbabwe, or across demographic, socio-economic, and behavioral factors that contribute to health disparities [6]. It is vital to monitor potential gaps in 95-95-95 progress, as populations lagging in these indicators are likely to be those in which HIV transmission and burden will concentrate in the future.

Geospatial analysis can help identify regions and communities that are lagging behind in each stage of the HIV care continuum, highlighting gaps in treatment and indicating the nature and location of programmatic response needed to close gaps in 95-95-95 [7]. Recent research has highlighted substantial spatial variability of HIV prevalence, incidence, and associated factors in Zimbabwe and elsewhere in sub-Saharan Africa, with variations in distribution at the province, district, and even finer scales [8-10]. However, no studies to our knowledge have examined spatial variability and factors associated with disparities in 95-95-95 targets in particular in Zimbabwe.

Against this background, the present study aimed to evaluate the progress towards achieving the 95-95-95 targets in Zimbabwe and identify factors associated with disparities in each of the three 95s. This analysis could help to inform HIV program planning, particularly resource allocation and intervention strategies tailored to meet the unique needs of specific geographic areas and populations.

## 2 Methods

### 2.1 Data sources

The primary data source for this study was the most recent publicly available 2015-2016 Zimbabwe Population-based HIV Impact Assessment (ZIMPHIA) [3]. The PHIA surveys are nationally representative household-based surveys of HIV status, risk factors, and care coverage funded by the United States (U.S.) President’s Emergency Plan for AIDS Relief (PEPFAR). Government of Zimbabwe conducted the ZIMPHIA from October 2015 to August 2016 through the Ministry of Health and Child Care (MOHCC), assisted by U.S. Centers for Disease Control and Prevention (CDC). ICAP at Columbia University conducted the study in association with Westat and other local partners. Using a two-staged stratified cluster sample design, ZIMPHIA identified 500 enumeration areas (EAs) based on probability proportional to size method in first stage, and households within EAs in second stage. Persons aged 15 years old and older living in a household or visitors who slept at that household the night before the survey were eligible for this survey with consent. We considered only adults aged 15 and older in this research. The survey offered home-based HIV testing and counseling (HBTC) with same day return of results, measured VLS and collected information on participating HIV treatment system, selected behaviors, sociodemographic factors through face-to-face interviews and HIV biomarkers. At the time of submission of this work, the subsequent (2020) ZIMPHIA survey dataset was still under internal analysis by the survey team and requests for access by external researchers were unable to be granted. Accordingly, our methodological approach was designed to apply to subsequent ZIMPHIA survey data as they become available.

HIV status in ZIMPHIA was determined using the Zimbabwean national HIV rapid-test algorithm. Individuals with a nonreactive screening test (Determine™ HIV-1/2 [Abbott Molecular Inc., Des Plaines, Illinois, United States]) were classified as HIV negative. Individuals with a reactive screening test result underwent a confirmatory test (First Response® HIV 1-2.O Card Test (Premier Medical Corporation Ltd., Nani Daman, India) and reported as HIV positive with a reactive confirmatory test. Individuals with a reactive screening test, followed by a nonreactive confirmatory test result, underwent a tiebreaker test (HIV 1/2 Stat-Pak™ [Chembio Diagnostic Systems, Medford, New York, United States]). Individuals with a reactive tiebreaker test were identified as HIV positive and those who received a nonreactive tiebreaker test result were identified as HIV negative.

### 2.2 Main outcomes

HIV-positive adults were classified as unaware of their HIV status if self-reported HIV status was reported as negative or unknown, and if none of three antiviral drugs tested in ZIMPHIA blood specimens (efavirenz, lopinavir, and nevirapine) were present above their respective limits of detection. Alternatively, they were classified as aware of status if self-reporting positive status or having detectable antiviral drugs. Individuals aware of their HIV-positive status were considered to be on ART if they reported current use of ART or ARVs were detected in their blood. Individuals on ART were considered to have VLS if the measured viral load (VL) in ZIMPHIA blood specimens was less than 1,000 HIV RNA copies/mL of plasma.

### 2.3 Spatial analysis

ZIMPHIA provided the geographical coordinates of the 500 EAs, which were recorded using the global positioning system (GPS) during the household survey. We used the kernel interpolation method to spatially smooth the HIV prevalence and estimates of each of the HIV care continuum measures: HIV status awareness, ART status, VLS, and HIV prevalence for each EA. We aggregated the spatially smoothed averages by district level to visualize the distribution of the estimates at the district level [11-13]. We performed optimized hotspot analysis [14] to identify the geospatial structure of the prevalence of HIV along with the prevalence of the three UNAIDS targets, with the identification of hotspots (areas with high HIV prevalence and high percentages of HIV awareness, ART uptake, or VLS) and coldspots (areas with low HIV prevalence and low percentages of HIV awareness, ART uptake, or VLS). Confidence intervals for hotspot analysis were calculated using Gi* statistics p-value in ArcGIS pro 2.8.

The geospatial structure of the HIV care continuum in Zimbabwe was also analyzed using spatial multivariate analysis in the geospatial GeoDa environment [11, 13, 15]. Multivariable spatial associations between all three metrics, HIV status awareness, ART status, and VL suppression, in each district were estimated using K-means clustering analysis. K-means is a partitioning clustering method in which the data are partitioned into k groups (i.e., fourth groups). In this clustering method, the n observations are grouped into k clusters such that the intra-cluster similarity is maximized (or dissimilarity minimized), and the between-cluster similarity minimized (or dissimilarity maximized). A further detailed description of these geospatial methods can be found elsewhere [16-18]. Percentages for the metrics, HIV status awareness, ART status, and VL suppression in each cluster identified were reported.

Lastly, we conducted a comparison between different vulnerability indexes for the health system, transport availability and housing, and socioeconomic factors in each cluster identified. These indexes were obtained from the Africa Covid Community Vulnerability Index from Surgo Ventures (https://precisionforcovid.org/africa) [19], and they measure several factors that assess the community vulnerability of determinants like the health system (health system strength, health system capacity, and access to health care), access to housing and transportation (household crowding, improved housing, sanitation, access to transportation, and road connectivity), and socioeconomic factors (access to information, education, poverty, and unemployment). We used ArcGIS pro 2.8 to generate the results and maps of percentages and clusters of HIV prevalence, HIV status awareness, ART status, and VLS.

### 2.4 Behavioral and sociodemographic covariates

We considered some sociodemographic and behavioral covariates to identify the constraints towards the achievement of 95-95-95 targets. The variables were education (no education/completed primary, completed secondary), wealth index (lowest, second/middle, fourth/highest), religion (Christians, no religion, other), age of the first sex contact (<15, 15-24, >=25), marital status (married/living together, divorced/separated/widowed, never married), place of residence (urban, rural), working status in last 12 months (work, did not work), and away from home in last 12 months (away from home, not away from home).

### 2.5 Statistical analysis

We estimated each of the metrics of HIV care continuum by sex and age groups using svymean() function from R survey package, and visualized using GraphPad software. Estimates incorporated survey weights to the ZIMPHIA’s multistage sampling procedure [20]. We conducted three distinct survey-weighted multivariate logistic regression models to assess the association between the selected sociodemographic and behavioral covariates and the three main outcomes of the study, HIV status awareness, ART and VLS. For Model 1: HIV status awareness, the survey data was subsampled to include HIV-positive individuals only; for Model 2: ART status, the survey data was subsampled to include HIV-positive individuals aware of their status only; and for Model 3: VLS, the survey data was subsampled to include HIV-positive individuals aware of their status and on ART treatment. We used svyglm() function for fitting the survey-weighted multivariate logistic regression models following two-stage sampling design of ZIMPHIA. From the survey data we created targeted subsamples for the three models using the svydesign() and subset functions.

### 2.6 Ethical considerations

The PHIAs were funded by PEPFAR with technical assistance through the US Centers for Disease Control and Prevention (CDC) under the terms of the cooperative agreement U2GGH001226. The study protocol and all data collection tools were approved by the Medical Research Council of Zimbabwe (A/1914) and the Institutional Review Boards at the U.S. Centers for Disease Control and Prevention (CDC) (IRB 6702), Westat (IRB 6317), and Columbia University (IRB Y06M00). This study follows the Strengthening the Reporting of Observational Studies in Epidemiology (STROBE) reporting guideline.

### 2.7 Data availability statement

Data are available in a public, open-access repository. The data that support the findings of this study are available from the Population-Based HIV Impact Assessment (PHIA; https://phiadata.icap.columbia.edu/), but restrictions apply to the availability of these data, which were used under license for the current study and so are not publicly available. We sought and were granted permission to use the core data set for this analysis by PHIA.

### 2.8 Funding

Research reported in this publication was supported by the National Institute of Mental Health and the National Institute of Allergy and Infectious Diseases of the National Institutes of Health under award numbers 5R01MH124478 and 1R01AI174932. The content is solely the responsibility of the authors and does not necessarily represent the official views of the National Institutes of Health.

## 3 RESULTS

## 3.1 Demographics of HIV prevalence and care

Estimated HIV prevalence in 2016 was 14.1% (95% confidence interval [CI]: 13.6% - 14.6%) of adults aged 15+, 12.2% (95% CI: 11.5% - 12.9%) for men and 15.9% (95% CI: 15.3% - 16.5%) for women. Across age groups included in the analysis, the highest HIV prevalence was observed in ages 45 to 54 at 27.8% (95% CI: 25.9% - 29.7%), while the lowest HIV prevalence was observed in ages 15 to 24 at 4.6% (95% CI: 4.1% - 5.1%). Among men, HIV prevalence was highest in ages 45 to 54 at 29.2% (95% CI: 26.1% - 32.3%). Among women, HIV prevalence was highest in ages 35 to 44 at 29.1% (95% CI: 27.2% - 31.0%). HIV prevalence was similar in urban areas (15.1%, 95% CI: 14.2% - 16.0%) and rural areas (13.6%, 95% CI: 13.1% - 14.1%). Across all of Zimbabwe, 78% (95% CI: 76.4% - 79.5%) of adults living with HIV were aware of their HIV status, 88.8% (95% CI: 87.5% - 90.1%) of those aware of their status were receiving ART, and 85.4% (95% CI: 83.9% – 87.0%) of those receiving ART were achieving VLS. Young women under age 25 were the least likely to be aware of their status, whereas young men ages 25-34, once aware of their status, were the least likely to be on ART, and young men under age 25 were the least likely to achieve VLS once on ART (Figure S1).

## 3.2 Geospatial analysis

Provinces like Matabeleland North, Matabeleland South, and Bulawayo had higher HIV prevalence than the national average with more than 18% HIV prevalence. More than 24% of the HIV-positive individuals were unaware of their HIV status in regions like Mashonaland West and the southern part of Masvingo. Likewise, more than 15% of people aware of their HIV status were not on ART in the northern parts of Matabeleland North and Midlands, and the southern part of Matabeleland South. In Mashonaland West, northern parts of Mashonaland Central, Mashonaland East, Manicaland, and southeast parts of Manicaland and not virally suppressed (Figure 2).

**Figure 1:**
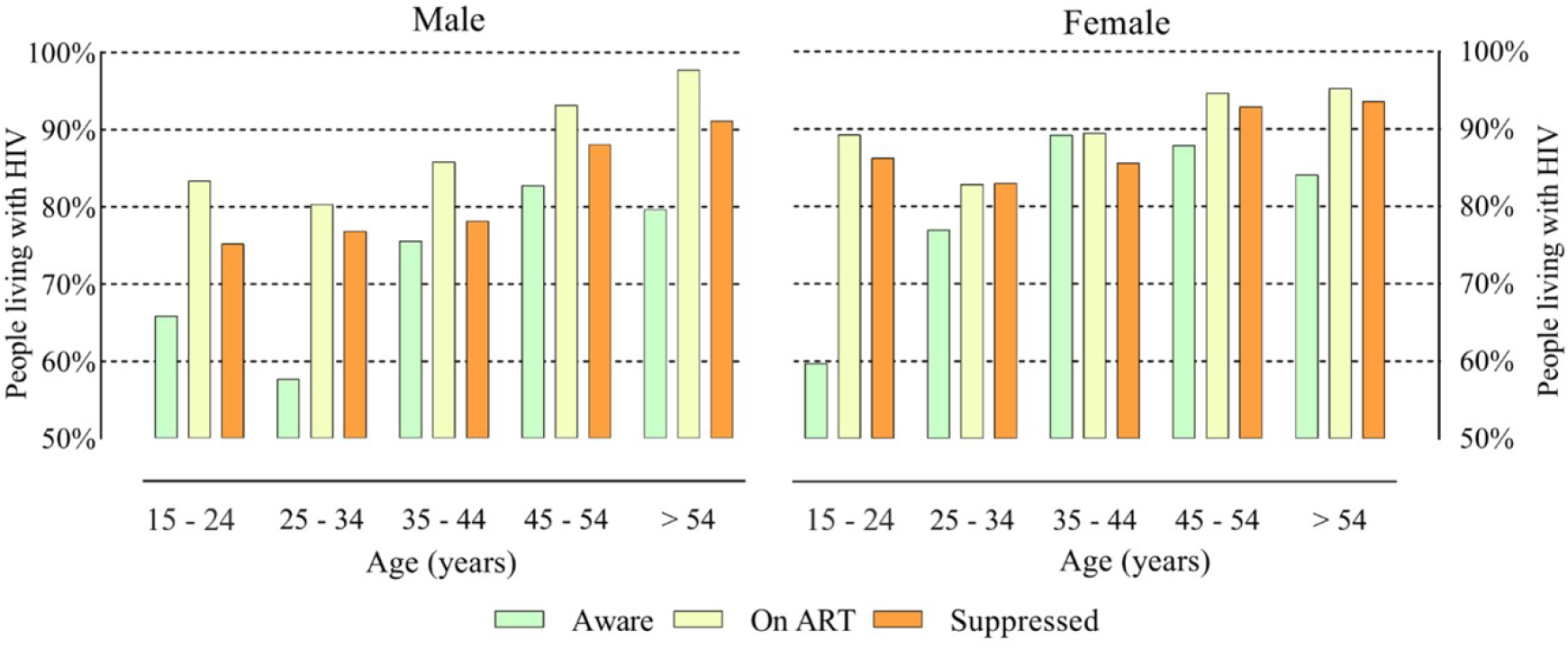
Demographic estimates of the 95-95-95 targets. Percentages of male and female categorized by age who are aware of their status, on ART and virally suppressed

**Figure 2:**
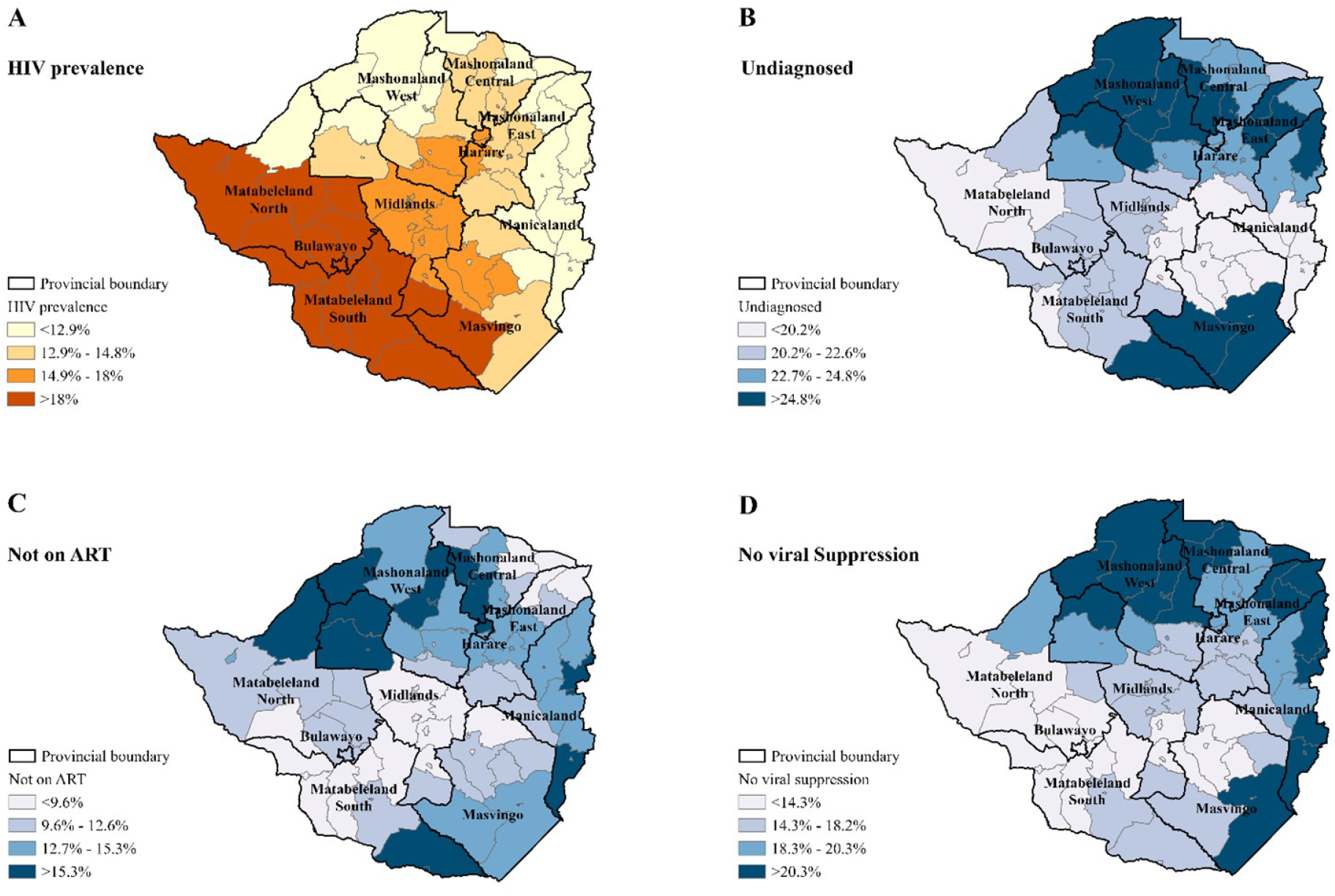
Maps of HIV prevalence and 95-95-95 targets in Zimbabwe. District level maps with percentage of (A) HIV prevalence, (B) HIV positive population who are unaware of their status, (C) people who are aware of their status but not on antiretroviral therapy (ART), (D) people who are on ART but not virally suppressed. Maps were created using ArcGIS Pro by ESRI version 2.8 (http://www.esri.com)

Masvingo more than 20% of people on ART were Hotspot analysis identified spatial clusters of high HIV prevalence in the southern part of the country, between the provinces of Matabeleland North and South, and Bulawayo (Figure 3A). Conversely, HIV-positive individuals unaware of their HIV status were clustered in the northern part of the country, between the provinces of Mashonaland West, Central, East, and Harare (Figure 3B). Individuals aware of their HIV status but were not on ART clustered in northern parts of Matabeleland North, and Mashonaland West provinces (Figure 3C). Furthermore, spatial clusters of individuals on ART but not virally suppressed were in Mashonaland Central, West and East, and Harare provinces (Figure 3D).

**Figure 3:**
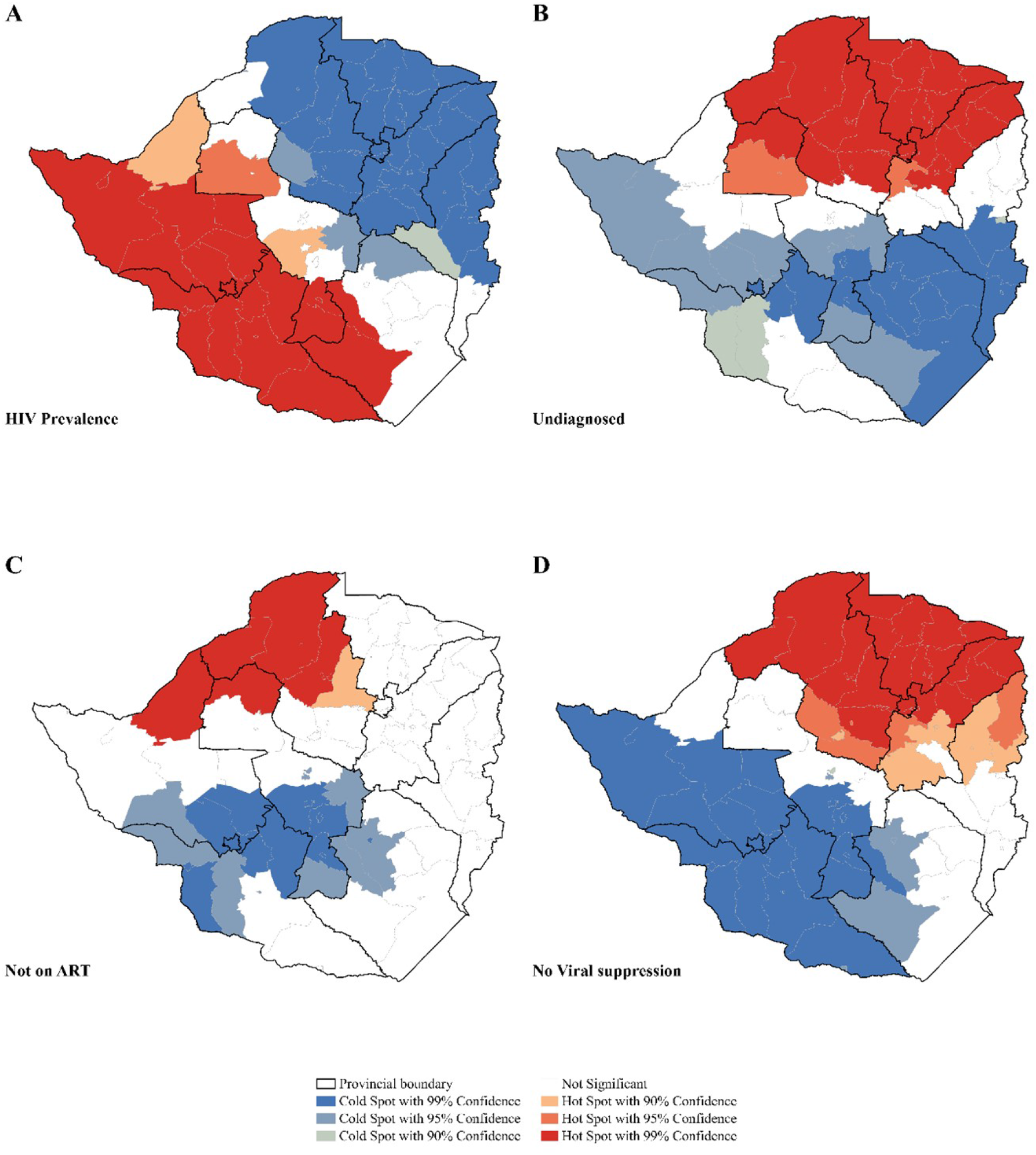
Hotspot maps of HIV prevalence and 95-95-95 targets in Zimbabwe. Hotspot and coldspot clusters of (A) HIV prevalence, (B) HIV positive population who are unaware of their status, (C) people who are aware of their status but not on antiretroviral therapy (ART), (D) people who are on ART but not virally suppressed

K-means clustering analysis identified four multivariable clusters of the different HIV care continuum metrics (Figure 4). Cluster 1 was mainly located in low HIV prevalence areas, with 13.5% HIV prevalence. In this cluster 24.3% of the HIV-positive people were unaware of their HIV status, 13.48% PLHIV aware of their status were not on ART, and 20.0% of HIV-positive individuals on ART were not virally suppressed. Cluster 2, located in the high HIV prevalence area with an estimated 19.8% HIV prevalence, had also 21.1% of the PLHIV unaware of their status, 9.8% of the HIV-positive individuals aware of their status were not on ART and 12.5 % of them were not virally suppressed. In Cluster 3, HIV prevalence was 13.6%, and among these HIV-positive individuals 18.6% were unaware of their status, 12.6% of these individuals were not on ART and 17.4% of them were not virally suppressed. The lowest HIV prevalence among these clusters was identified in Cluster 4, with an estimated 11.6% HIV prevalence. Among the HIV-positive individuals in this cluster, 26.8% were unaware of their status, 21.3% of those aware of their status were not on ART, and 29.4% HIV positive individuals on ART were not virally suppressed.

**Figure 4:**
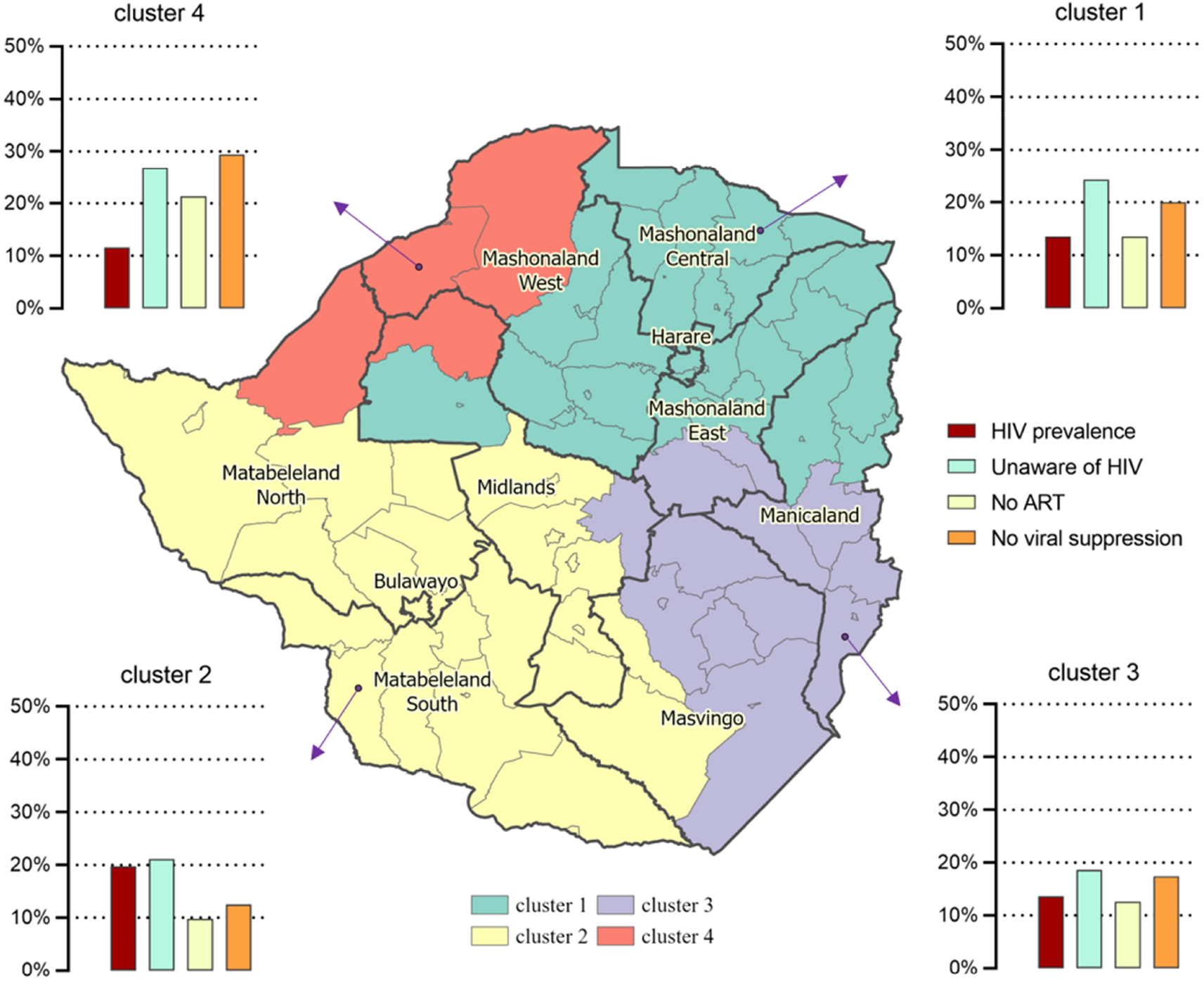
K-means cluster map showing clusters of HIV prevalence and care continuum patterns. This map provides clusters combining HIV prevalence, HIV positive population who are unaware of their status, people who are aware of their status but not on antiretroviral therapy (ART), people who are on ART but not virally suppressed

The health index combined with the ART uptake and viral suppression maps indicated that both, the health index and ART uptake were high in Mashonaland East province. The health index in Mashonaland West and Manicaland provinces was high, but ART uptake was low. In Harare, both the health index and ART uptake were low. Likewise, viral suppression in Mashonaland West and Manicaland provinces was low, but the health index was high (Figure 5).

**Figure 5:**
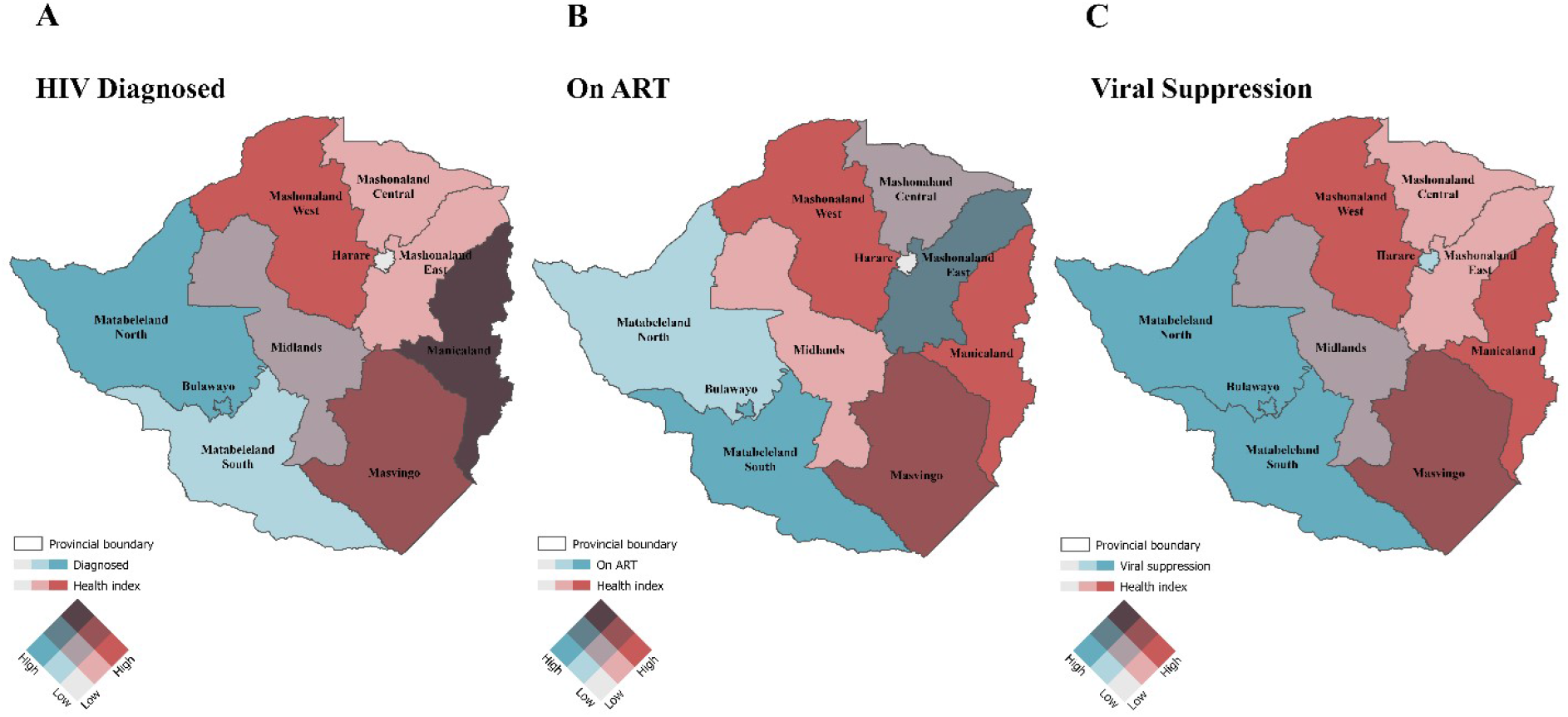
Bivariate maps of health index and UNAIDS 95-95-95 targets. Maps of health index with (A) HIV positive population who are aware of their HIV status, (B) people who are aware of their status and on antiretroviral therapy (ART), (C) people who are on ART and virally suppressed. Maps were created using ArcGIS Pro by ESRI version 2.8 (http://www.esri.com)

## 3.3 Factors associated with care continuum gaps

At the individual level, care continuum gaps were more common in men and less common in older age groups (Figure 6). Employment and belonging to a minority religion or no religion were associated with lack of diagnosis (Figure 6A), but not with gaps in ART (Figure 6B) or VLS among those diagnosed (Figure 6C). Other factors (education, wealth, age at first sex, marital status, urban/rural residence, and travel away from home) were not significantly associated with care continuum gaps in multivariable analysis

**Figure 6:**
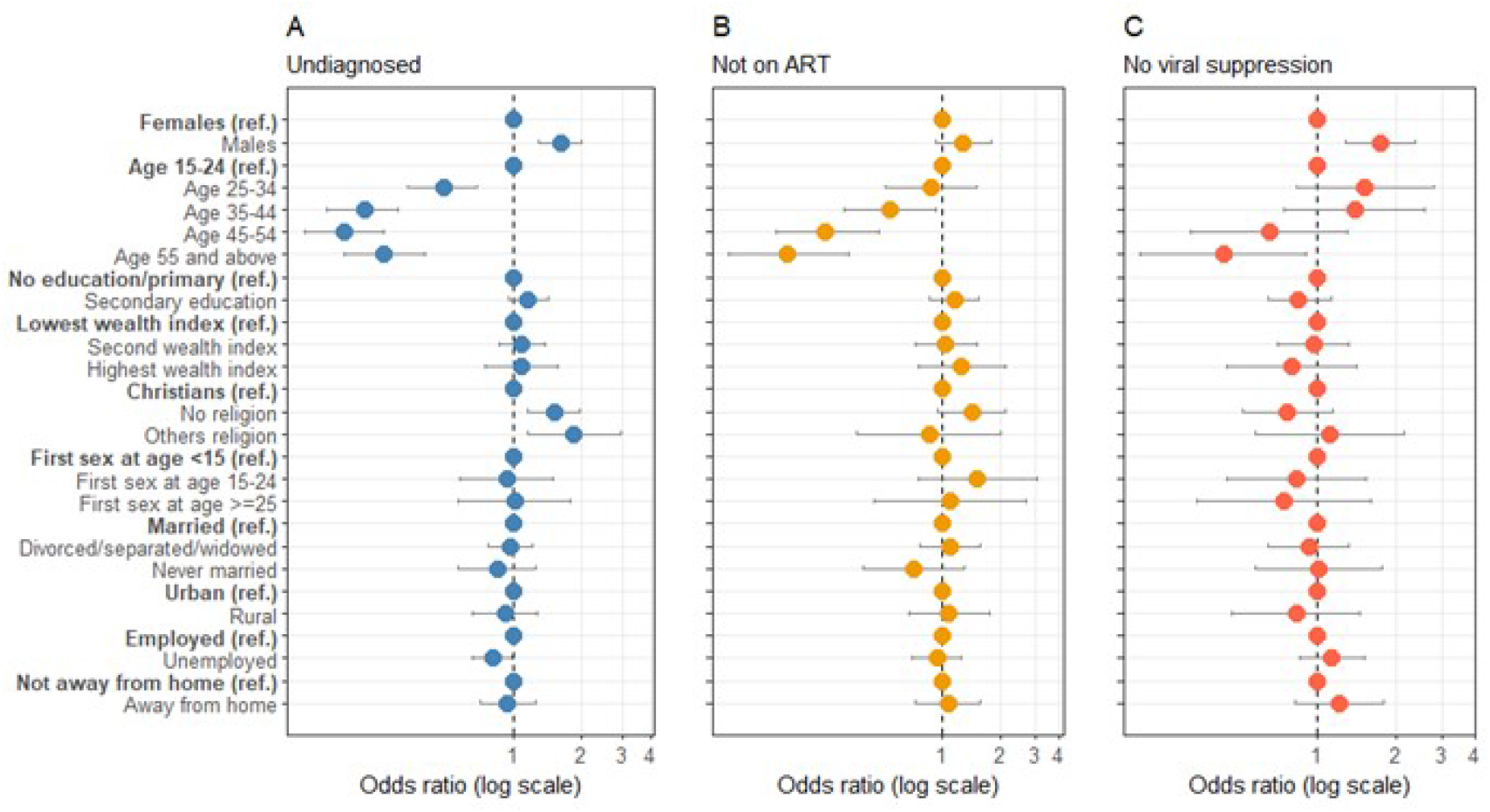
Multivariable logistic regression models. Odds ratio of socio-demographic and behavioral factors associated with HIV-positive people (A) who are unaware of their HIV status -Model 1, (B) who are aware of their status but not on antiretroviral therapy (ART) -Model 2, (C) who are on ART but not virally suppressed -Model 3

## 4 DISCUSSION

The Republic of Zimbabwe has made significant strides towards meeting the ambitious UNAIDS 95-95-95 targets, which aim to achieve a 95% reduction in new HIV infections, 95% access to ART, and 95% viral suppression by 2030. Despite these positive health outcomes, regional heterogeneity in the different UNAIDS 95-95-95 targets was observed in this study. Spatial analysis revealed contrasting patterns between the HIV prevalence and the UNAIDS 95-95-95 targets. Specifically, high HIV prevalence was mostly concentrated in the southern part of the country, primarily between the provinces of Matabeleland North and South. Conversely, poor UNAIDS 95-95-95 target outcomes were mostly concentrated in the northern part of the country, including the provinces of Mashonaland West, Central and East, and Harare. These findings have important implications for decision-making and resource allocation, as targeted interventions tailored to the specific needs of these regions and vulnerable communities could help to improve UNAIDS 95-95-95 target outcomes and ultimately advance progress towards ending the HIV epidemic.

In the context of several African countries, including Zimbabwe, HIV testing facilities are predominantly located in areas with high HIV prevalence, where HIV screening is mandatory to curb the epidemic in these high HIV burden areas [3, 21]. Consequently, PLHIV in low HIV prevalence areas may face limited access to HIV testing facilities, resulting in an increased number of PLHIV unaware of their status in these areas. In this study, we found that HIV-positive individuals who were unaware of their status were primarily concentrated in areas with low HIV prevalence, while those from high HIV prevalence areas were mostly aware of their HIV status. Additionally, areas with low HIV prevalence had higher percentages of PLHIV who were not on ART and those on ART but were not virally suppressed. Our multivariate clustering analysis revealed an area with the slowest progress in achieving 95-95-95 (Cluster 4), characterized by the highest percentages of PLHIV who were unaware of their HIV status, those who were aware of their HIV status but not on ART, and those who were on ART but not virally suppressed. These findings highlight the need to expand HIV testing and treatment services beyond areas of high HIV prevalence to better address regional disparities in HIV care continuum outcomes.

Our study identified a concentration of areas with high HIV prevalence and poor progress towards achieving the UNAIDS 95-95-95 targets in border regions with high levels of migrant labor and population mobility. Migrant workers from neighboring countries, such as Malawi, Mozambique, and Zambia, encounter significant difficulties in accessing adequate healthcare facilities and routine HIV treatment services, which are vital for HIV diagnosis and control [22, 23]. Despite the relatively higher health index in these regions, progress towards achieving the UNAIDS 95-95-95 targets has been slower, which may be due to low HIV per capita spending. The two primary metropolitan provinces in Zimbabwe, Harare and Bulawayo, both have a low health index. However, there are significant differences in ART coverage and viral suppression attainment between these provinces, which may be due to variations in per capita HIV spending. Bulawayo, which spent twice as much as Harare in 2012, may have made better progress in HIV treatment [24]. Conversely, provinces such as Mashonaland West have low ART uptake and viral suppression rates despite having a better health index, possibly due to the lowest per capita HIV spending in 2012 [24]. Males had a lower level of HIV status awareness and were less likely to achieve viral suppression compared to females, which is consistent with other studies conducted in Zimbabwe, Italy, Venezuela, Cameroon, and Central Haiti, using nationally representative and clinical case study data [25-29]. In all studies, males were identified as latecomers to HIV/AIDS care. Moreover, we found that younger individuals with HIV had lower odds of being aware of their HIV status, receiving ART, and achieving viral suppression, while older individuals were more likely to be tested for HIV, more inclined to receive ART and routine checkup, and had better viral control with lower viral loads [30-33].

Achieving the UNAIDS’s ambitious 95-95-95 targets in resource-constrained countries like Zimbabwe requires a well-designed plan that targets high-risk and underserved communities with a high burden of infection and slow progress towards these targets [34]. However, providing HIV treatment facilities to everyone on a national basis might not be feasible, and thus, intervention programs should be implemented at the sub-national and local level. In this regard, it is critical to identify communities with low first 95 prevalence areas and ensure regular access to healthcare and HIV testing by decentralizing HIV treatment services in Zimbabwe, thereby overcoming barriers to linkage to care. It is important to note that sociocultural behaviors associated with HIV, healthcare access, and funding for HIV treatment vary across local communities, leading to spatial heterogeneity of HIV prevalence and the distribution of the 95-95-95 targets with distinctive clusters as showing in this study [35]. Therefore, tailored HIV testing and treatment programs need to be implemented independently to balance the spatial disparity of progress towards the UNAIDS 95-95-95 targets, ultimately addressing the spatial heterogeneity for each of the targets observed in the country.

Likewise, the impact of migration and mobility on the spread of HIV in African nations has been widely acknowledged, and therefore, ongoing intervention initiatives must include migrant workers from border regions [36]. These workers face challenges such as a lack of stable habitats and limited access to treatment, highlighting the need for a system that can link them to treatment, which is critical for achieving viral suppression. Furthermore, improving the involvement of migrant workers in HIV treatment facilities is essential, as it becomes increasingly difficult to attain viral suppression after an extended period of HIV infection. It would be recommended that areas with high HIV prevalence and low progress towards UNAIDS 95-95-95 targets should be provided with additional funding for HIV treatment. Provinces such as Mashonaland West and Harare, which have low per capita HIV spending, require increased funding to address the HIV epidemic. In 2012, the majority of HIV funds in Zimbabwe were spent at the national level, which may not have helped provinces with low progress towards achieving the UNAIDS 95-95-95 targets [24]. Instead, it may be more effective to allocate funds at the subnational and local levels, considering the unique strategies required in each community. To eradicate HIV from an area, financial resources are necessary to provide better services, improve infrastructure, and conduct continuous monitoring of PLHIV. Additionally, community-based HIV testing may be a viable alternative to testing in healthcare facilities, as it is often better received in the community. By increasing testing in the community, individuals who are not infected can also be reached and educated on HIV and initial treatments [37].

### 4.1 Limitations

Our study has several limitations that need to be considered when interpreting the results. Firstly, we used data from a cross-sectional population survey, namely the PHIA survey, which may not represent the entire population. For instance, some high-risk subpopulations such as female sex workers, injection drug users, men who have sex with men, and migratory people may have been underrepresented due to absence or non-participation [38, 39]. Additionally, self-reported data may have been subject to recall errors or false information by respondents who may not have been aware of the survey [38]. Furthermore, because Zimbabwe shares borders with South Africa, Botswana, Zambia, and Mozambique, estimating the population in border areas can be challenging. Migrant people, in particular, are difficult to capture due to their dynamic nature, which may lead to an underestimation or double counting of their numbers, potentially affecting the study findings.

Although ART uptake and viral suppression rates are greater than HIV diagnosis, it should be noted that the cascade from PLHIV is not homogenous as the percentage of people unaware of their HIV status is high, it may result in lower estimates for other cascade indicators. Additionally, the number of people on ART may not be an accurate measurement, as the PHIA survey does not capture the regularity or adherence to ART. Finally, the spatial displacement procedure of households to protect the confidentiality of respondents may have also affected the estimates at the sub-national level. This is because households may have been displaced a few kilometers away from their actual location, leading to changes in the estimates. Therefore, these limitations should be considered when inter-10preting the findings of our study.

## 5 CONCLUSIONS

Our study highlights the necessity of localized targeted interventions that address sociodemographic constraints and geographic heterogeneity in Zimbabwe to achieve the UNAIDS’s 95-95-95 targets. These interventions could involve improving access to testing and treatment services in areas with lower coverage identified in our study, increasing education and awareness regarding HIV prevention and treatment, and overcoming social and cultural obstacles to healthcare service access. Moreover, our study emphasizes the significance of geospatial mapping tools to uncover communities with insufficient HIV care coverage and to design tailored interventions for those specific areas. In general, our study underscores the importance of a comprehensive and focused approach to meeting HIV/AIDS goals in Zimbabwe. This approach could serve as a model for other countries confronting similar challenges as they strive to achieve the 95-95-95 targets. It is crucial to prioritize and invest in effective strategies that address the sociodemographic constraints and geographic heterogeneity identified in this study to ensure equitable access to HIV prevention and treatment services for all individuals in Zimbabwe. By doing so, significant progress can be made toward achieving the 95-95-95 targets and ultimately working toward ending the HIV/AIDS epidemic as a public health threat by 2030, in line with the global Sustainable Development Goals. Further research is necessary in these low-progress areas to gain a better understanding of how to achieve the targets within the desired timeframe and to ensure that no one is left behind.

